# Obsessive-compulsive disorder is characterised by metacognitive deficits resistant to subthalamic stimulation

**DOI:** 10.1101/2024.11.03.24316467

**Authors:** Audrey Kist, Michael Pereira, Ramla Msheik, Dorian Goueytes, Arnaud Pouchon, Stephan Chabardès, Julien Bastin, Mircea Polosan, Nathan Faivre

**Affiliations:** Univ. Grenoble Alpes, Univ. Savoie Mont Blanc, CNRS, LPNC, 38000 Grenoble, France; Univ. Grenoble Alpes, Inserm, U1216, CHU Grenoble Alpes, Grenoble Institut Neurosciences, 38000 Grenoble, France; Department of Psychiatry and Behavioral Sciences, UCSF, San Francisco, USA; Department of Psychiatry, CHU Grenoble Alpes, F-38000 Grenoble, France; Univ. Grenoble Alpes, CEA, LETI, Clinatec, 38000, Grenoble, France; Univ. Grenoble Alpes, Inserm, Department of Neurosurgery, 38000, Grenoble, France

**Author notes:** equal contributions (Mircea Polosan, Nathan Faivre). equal contributions (Audrey Kist, Michael Pereira). **Corresponding author** Nathan Faivre, Laboratoire de Psychologie et Neurocognition, CNRS UMR 5105, UGA BSHM, 1251 Avenue Centrale, 38058 Grenoble Cedex 9.

**Keywords:** perceptual decision-making, metacognition, deep brain stimulation

## Abstract

Most human studies on perceptual decision-making are correlational, and few addressed the causal role of the neural markers they identify. Here, we took the opportunity offered by deep brain stimulation to study the causal role of subthalamo-cortical networks on perceptual decision-making and perceptual monitoring. To this end, we asked participants with obsessive-compulsive disorder (OCD) treated with subthalamic stimulation to detect visual stimuli and provide confidence reports while we recorded scalp electroencephalography. Following a preregistered plan, we compared behavioural and neural results between patients off-stimulation and matched healthy controls and among patients on and off-stimulation to identify the effect of subthalamic stimulation on perceptual choices and metacognitive judgments. Our results indicate that patients off-stimulation detected visual stimuli similarly to matched healthy controls but provided poorer confidence judgments indicative of a metacognitive deficit. Moreover, chronic subthalamic stimulation significantly reduced clinical symptoms, but acute stimulation changes did not impact visual detection, confidence, or corresponding event-related potentials at the scalp level. We conclude that acute changes in subthalamic stimulation are insufficient to alleviate metacognitive deficits observed in OCD.

## Introduction

Obsessive-compulsive disorder (OCD) is characterised by intrusive thoughts (obsessions) and irrepressible repetitive behaviours or mental acts (compulsions), responsible for severe effects on functioning and quality of life (Abramowitz et al., 2009). Individuals with OCD have a heightened sense of doubt (Abramowitz et al., 2009), with reduced confidence in their decisions based on perception or memory (Dar et al., 2022; Hoven et al., 2019), suggesting the existence of a metacognitive deficit. However, the physiopathological origins of these deficits remain poorly understood. Subcortical contributions to confidence are particularly relevant in OCD, as numerous studies point to a disruption of cortical-subcortical circuits in this disorder (Stein et al., 2019) and because chronic deep brain stimulation (DBS) of the subthalamic nucleus (STN) is now recognised as a promising treatment for drug-resistant OCD (Chabardes et al., 2020). During speeded discrimination tasks, the STN is assumed to modulate choices by modulating the threshold at which accumulated evidence leads to commitment to a decision (Cavanagh et al., 2011; Frank et al., 2007; Herz et al., 2016). This modulation can impact confidence, given that confidence is defined relative to the decision threshold, as assumed by numerous models based on signal detection theory and evidence accumulation (for reviews, see Mamassian, 2016; Mazancieux et al., 2023). Little is known regarding a possible subthalamo-cortical origin of metacognitive impairments in OCD, as most of the literature about the effects of STN stimulation concerns individuals with Parkinson’s disease and motor symptoms (Weintraub & Zaghloul, 2013).

Here, we sought to assess whether STN stimulation could modulate decision thresholds and alleviate metacognitive impairments in OCD. To this end, we asked individuals with OCD and control participants to detect visual stimuli and provide confidence judgments in their responses while we recorded scalp EEG. Following a randomised, double-blind procedure, patients with OCD were tested on and off subthalamic DBS. The following predictions were preregistered before data collection (https://osf.io/qcvsd). First, considering the perceptual and metacognitive deficits in OCD (Dar et al., 2022; Pushkarskaya & Ma, 2020), we expected higher detection thresholds and lower metacognitive performance among patients off-stimulation than control participants. Second, assuming that subthalamic DBS lowers perceptual thresholds (Cury et al., 2016; Voon et al., 2017), and that confidence is defined as the distance between sensory evidence and the threshold, we expected more liberal detection behaviour and higher confidence for seen stimuli and lower confidence for unseen stimuli in patients on-stimulation than off-stimulation.

## Materials and Methods

### Participants

We recruited individuals with OCD implanted with bilateral 8-lead directional electrodes (Boston Scientific, Marlborough, USA) in the anteromedial part of the STN at least 12 months before the study. Healthy participants with no current or past psychiatric or neurological conditions served as controls. They received 20€ as compensation for their time. Behavioural analyses were conducted on 14 participants with OCD (11 female; age = 44.1 ± 10.0 years; mean ± 95% confidence interval) and 18 healthy controls (13 female subjects; age = 45.5 ± 11.7 years). Supplementary Table 1 and Table 2 provide groups’ demographic information and clinical scores. The protocol was approved by an ethics committee (2012-A00490-43), and all participants provided written informed consent before the study.

### Procedure

Participants were exposed to a 4 s stream of visual noise frames, each presented for 200 ms. A face stimulus could be embedded within a frame at a pseudo-random time ranging from 0.6 to 3.6 s from trial onset. Stimulus intensity was manipulated by changing the contrast between the stimulus and the noise frame (0, 0.5, 0.75, 1, 1.25, or 1.5 times each participant’s detection threshold, measured before the experiment). After each trial, participants were asked to report whether they detected no face, one face, or more than one face. A randomly uniform delay between 0.5 and 0.75 s between the last noise frame and the report prompt ensured that EEG signals after the stimulus onset were not contaminated by motor activity due to the button press associated with the report. Next, participants reported their confidence in their detection report on a 3-level scale corresponding to: “not sure”, “moderately sure”, and “very sure”. The experiment lasted 1h maximum, corresponding to 5 to 7 blocks of 30 trials each. Participants in the OCD group were tested for two sessions, either with or without STN stimulation, in a randomised, double-blind way. Session order was counterbalanced across participants, and a washout period of 16 hours was respected between sessions (Fig. 1A-B).

**Figure 1.**
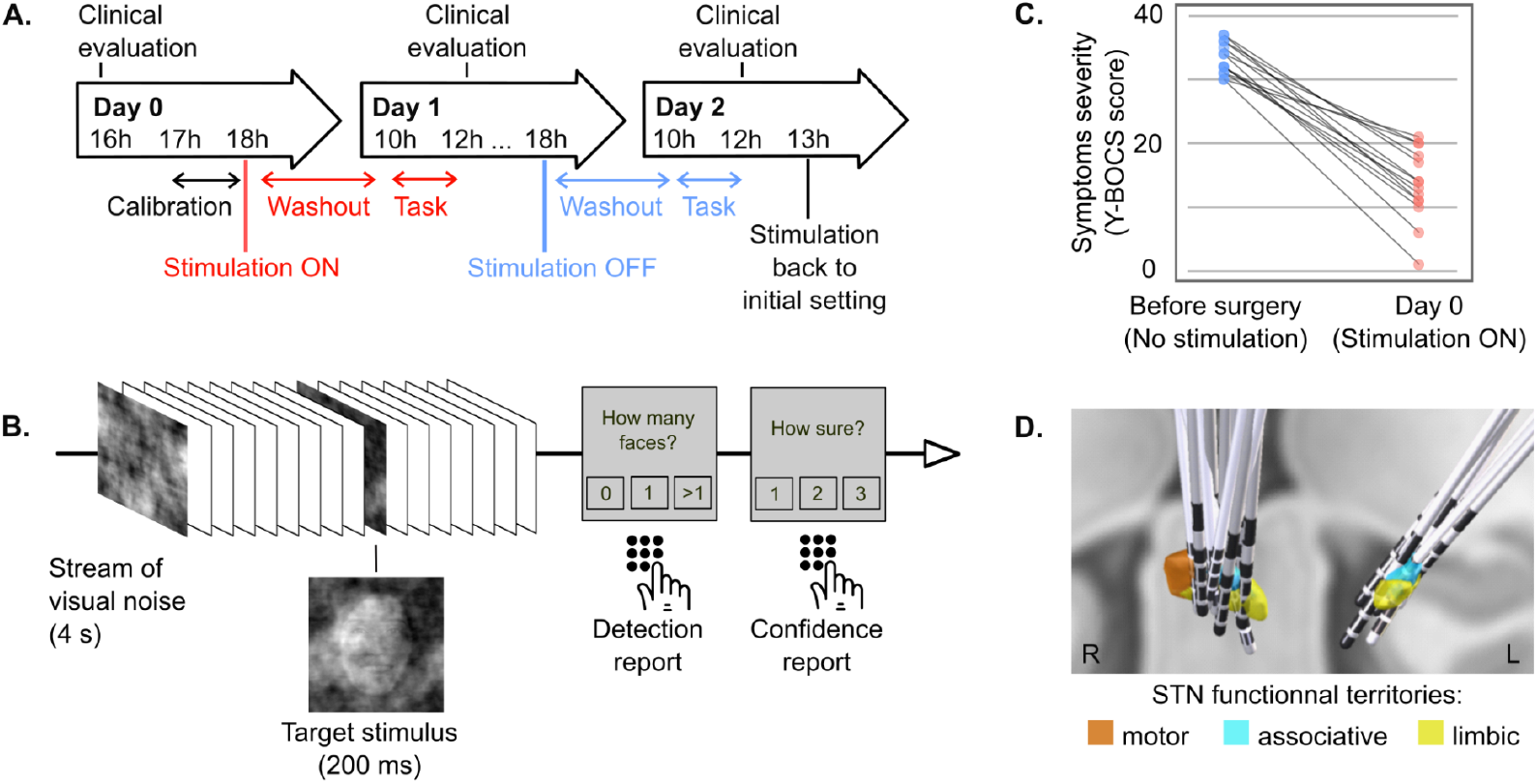
**A. Experimental procedure.** Participants in the OCD group were tested either with or without STN stimulation in a randomised, double-blind way. Session order was counterbalanced across participants. On day 0, a clinical evaluation was performed together with a presentation of the task, a detection threshold procedure, and a baseline behavioural assessment. Then, the DBS status was modified according to the double-blind randomisation. On day 1, the EEG and behaviour were recorded after a 16-hour washout, followed by a blind clinical evaluation. Depending on the randomisation, the DBS status was switched on or off in the late afternoon. The last recording happened on day 2, after a similar 16-hour washout period. After the recording, a final clinical evaluation was performed, and the DBS status was switched back to the initial on status. **B. Task**. A four-second stream of visual noise was presented. A 200 ms face stimulus could be embedded in this visual stream. The intensity level of the stimulus was adjusted to one of five intensity levels around the participant’s detection threshold (50%, 75%, 100%, 125%, 150% and catch trials). After four seconds, the participant reported the number of detected stimuli (0, 1, more than 1) and their confidence level (not sure, moderately sure, very sure) using a keyboard press. **C. Clinical scores before and after long-term subthalamic stimulation in participants with OCD**. In blue, the Y-BOCS score was measured at the time of the surgery, before the stimulation electrode implantation, i.e., without any subthalamic stimulation. In red, the Y-BOCS score was measured on day 0 of the protocol, with chronic subthalamic stimulation for at least one year. **D. Implantation of subthalamic electrodes in participants with OCD**. The three territories of the STN are represented in orange (motor territory), blue (associative territory), and yellow (limbic territory). The electrodes target the blue-yellow area, where most contacts are located.

### Behavioural analysis

We analysed detection and confidence as a function of stimulus intensity and group (controls, patients off-stimulation, patients on-stimulation) using Bayesian mixed-effects regressions (see SI). Detection reports (yes/no) were analysed with a Bayesian mixed-effects binomial regression with group (control vs off, on vs off), stimulus intensity (0, 0.5, 0.75, 1, 1.25, 1.50 detection threshold), and stimulus onset (0.6 to 3.6 s) as fixed effects:

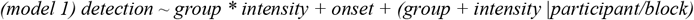

Confidence was analysed with a Bayesian mixed-effects ordinal regression with group, stimulus intensity, detection answer (yes/no), and stimulus onset as fixed effects:

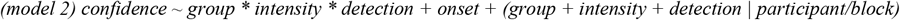

### Electroencephalography

EEG was acquired with 64 electrodes at a sampling rate of 5000 Hz. All the preprocessing steps were performed using the Matlab Fieldtrip toolbox and in-house scripts. The main steps were low-pass filtering (45 Hz), downsampling to 500 Hz, high-pass filtering (0.5 Hz), additional notch filters at 50 Hz and half the stimulation frequency, epoching around stimulus onset, re-referencing to a common average, and independent component analysis (see SI). We first identified clusters of responsive electrodes. An electrode was considered responsive when it presented any significant effect using the following models, during at least 50 ms in a row, adjusting for false discovery rate. The models were linear mixed-effects regressions for each electrode and each time sample, excluding catch trials in which no stimulus was presented:

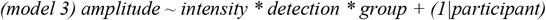

Model 4 was similar to model 3 but considered trials with detected stimuli only (HIT), and confidence instead of detection:

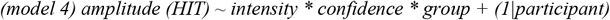

Effects of interest were further examined with Bayesian models, including a full random effect structure (see Rouy et al., 2023 for a similar approach). Namely, we selected the electrode with the most prolonged effect for each model. We formed a cluster with its surrounding electrodes showing significant effects for at least 50 ms at any time during this time window. Then, we averaged the amplitudes over electrodes and over the corresponding time window to fit the following models:

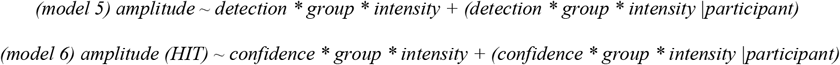

Categorical variables were dummy-coded, with the following references: control group or DBS-off group, low confidence, no stimulus detection.

## Results

### Metacognitive deficit in individuals with OCD

We first compared demographic and clinical data between control participants and patients with OCD at the beginning of the protocol, on-stimulation. The groups did not differ significantly in age, education, anxiety, or depression (SI Table 1). Patients had received chronic, continuous STN stimulation for at least one year (mean stimulation duration = 5.7 years ± 1.2 years, Fig. 1D), resulting in a significant reduction of symptoms score as measured on the Yale-Brown Obsessive Compulsive Scale (mean Y-BOCS before surgery = 33.36 ± 2.61, after surgery = 13.43 ± 5.82, BF > 8000, Fig. 1C). We next sought to assess if OCD was associated with altered perceptual abilities by comparing the probability of a stimulus being detected (stimulus detectability) between groups (control participants vs patients off-stimulation) as a function of sensory evidence (stimulus intensity). Contrary to what we had pre-registered, we found inconclusive evidence concerning the main effect of group (estimate −0.37 [−1.89 1.14]; BF = 0.38) and interaction between group and stimulus intensity (estimate 0.56 [−0.94 2.02], BF = 0.41, Fig. 2A-B, SI Table 4). These results suggest that participants with OCD off-stimulation and control participants had qualitatively similar perceptual performances. We also checked that there was no difference between the absolute value of the detection threshold measured in participants with OCD at day 0, compared to control participants (t[29.54]= −1.29, BF = 0.59). At the neural level, we found equivalent EEG responses between groups. A Bayesian mixed-effects regression (model 5) on a cluster of electrodes around CPz showing an interaction between detection and intensity revealed no conclusive difference between groups (Fig. 2C-D and SI Tables 5-6 for full model results and electrode selection). Together, these results indicate that participants with OCD showed no impairment in perceptual decision compared to control participants, making a direct comparison of confidence between the two groups possible.

**Figure 2.**
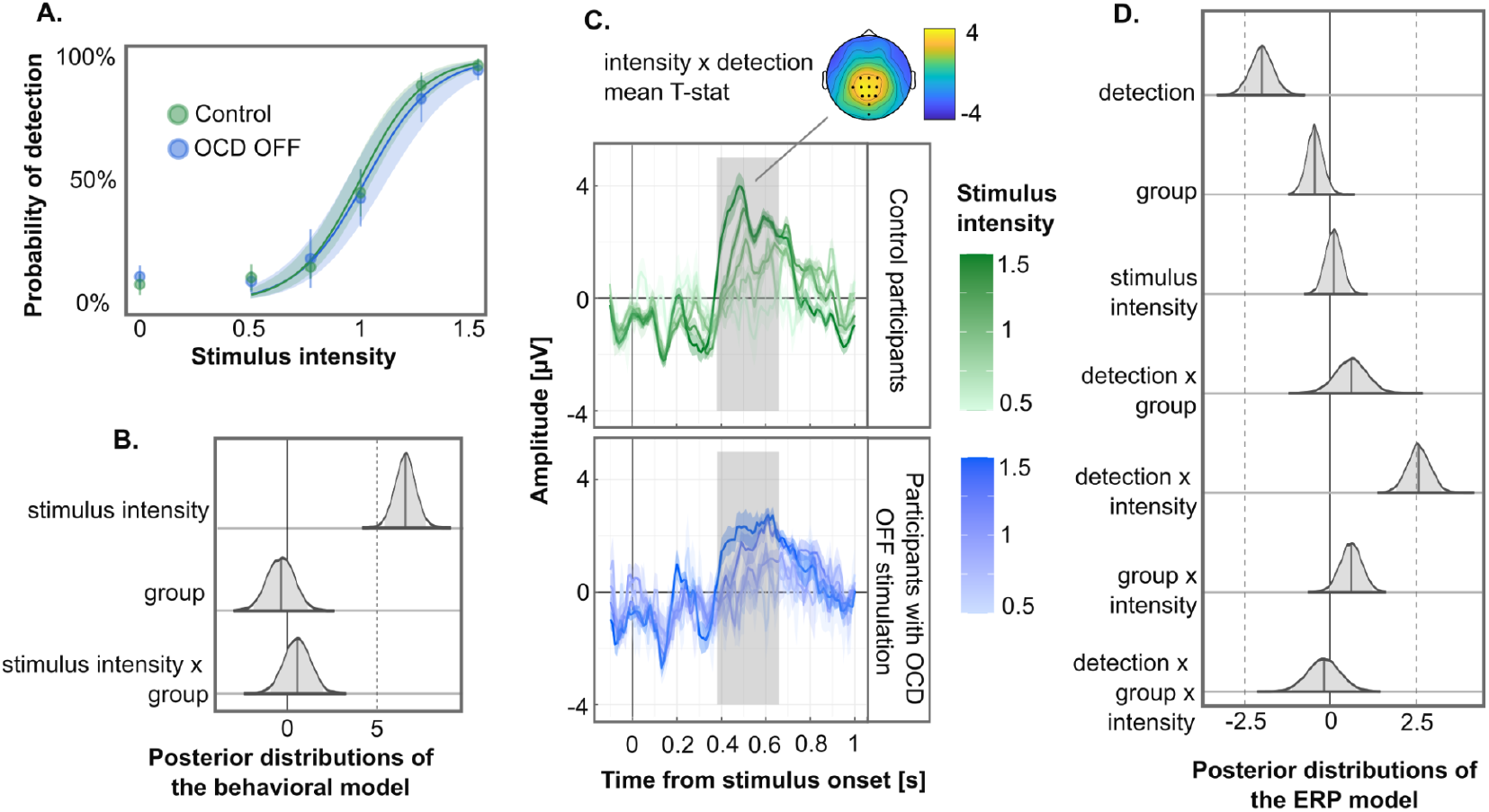
Participants with OCD off-stimulation (in blue) compared to control participants (in green). **A. Detection reports**. The dots show the mean probability of detection for each stimulus intensity level. The line and ribbon show the Bayesian model prediction and 95% confidence interval. **B. Posterior distributions from the Bayesian model predicting detection reports. C. Detection-related event-related potential (ERP)**. The topography represents the mean t-statistic of the most significant effect during the time window, i.e., mean effect of intensity, between 0.38 and 0.51 after stimulus presentation. The dots represent the electrodes of interest, i.e., electrodes responding to intensity for more than 50 ms in a row, within the time window of interest, and surrounding the most responsive electrode, Pz. Below is the amplitude of the evoked ERP on the cluster of interest. The time window of interest is shown in grey. **D. Posterior distributions from the Bayesian model predicting EEG amplitudes depending on detection reports**.

Contrary to stimulus detection, we found an interaction between group, response, and stimulus intensity on confidence (estimate = 0.90 [0.14 1.65], BF = 2.56). We interpret this as weak evidence as the credible interval did not overlap with zero, but BF was smaller than 3 (SI Table 10). This interaction implied that confidence tracked the intensity of detected stimuli less well in the OCD than in the control group, in line with a metacognitive deficit in the OCD group (Figure 3A-B). Time-wise analyses of EEG amplitude revealed an interaction between intensity and confidence over a cluster of electrodes around Pz (SI Table 10). Follow-up Bayesian analyses at the cluster level revealed an absence of a main effect of group (estimate = −0.1 [−1.38 1.15], BF = 0.30) and inconclusive evidence concerning the interaction between group and other independent variables (Fig. 3C-D, SI Table 11-12). Together, these results indicate that the metacognitive deficit observed behaviorally was not observable in terms of EEG amplitude at the scalp level.

**Figure 3.**
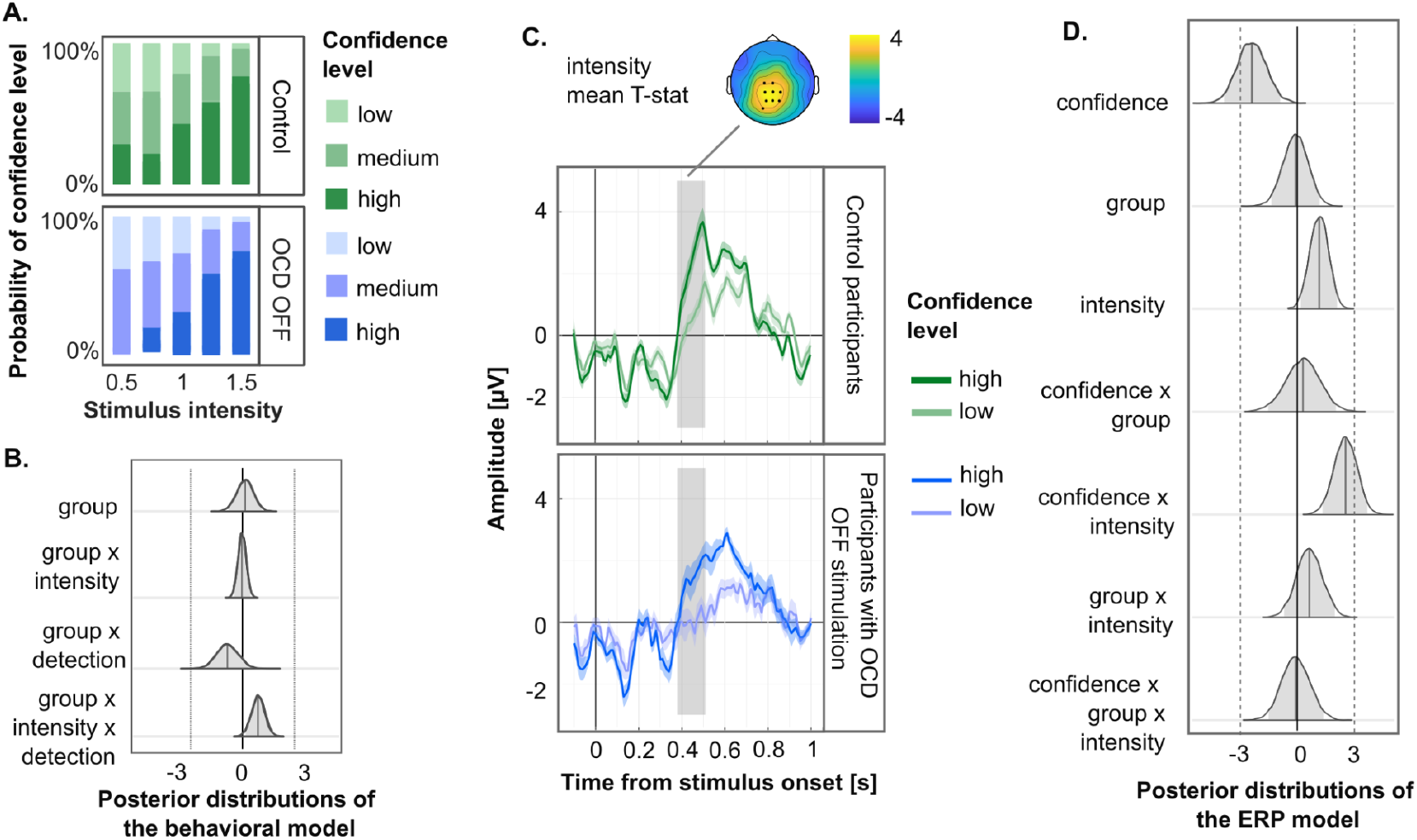
Participants with OCD off-stimulation (in blue) compared to control participants (in green). **A. Confidence reports**. Probability of confidence level for each stimulus intensity level. **B. Posterior distributions from the Bayesian model predicting confidence reports**. Only effects related to the group are shown here. **C. Confidence-related evoked potential**. The topography represents the mean t-statistic of the most significant effect during the time window, i.e., mean effect of intensity, between 0.38 and 0.51 after stimulus presentation. The dots represent the electrodes of interest, i.e., electrodes responding to intensity for more than 50 ms in a row, within the time window of interest, and surrounding the most responsive electrode, Pz. Below is the amplitude of the evoked ERP on the cluster of interest. The time window of interest is shown in grey. Low and medium-confidence reports have been pooled into a single Low level to increase plot readability, and all stimulus intensity levels are pooled together. **D. Posterior distributions from the Bayesian model predicting EEG amplitudes depending on confidence reports**.

### No effect of STN stimulation on detection and confidence

Next, we assessed the clinical effect of DBS by comparing clinical outcomes on and off-stimulation. OCD symptoms measured with the Y-BOCS were lower on-stimulation before the experiment (mean 13.43 ±5.42) than before surgery (mean 33.36 ± 2.31; t[13] = 12.69, BF > 8000). However, Y-BOCS sub-scores on- and off-stimulation collected after each experimental session did not differ significantly (SI table 2).

To evaluate the impact of STN stimulation on stimulus detectability, we compared detection reports on- and off-stimulation. We found evidence for an absence of a stimulation effect (estimate 0.34 [−0.54 1.21], BF = 0.25), and absence of interaction between stimulation and stimulus intensity (estimate = −0.04 [−0.80 0.72], BF = 0.18, Fig. 4 A-B, SI Table 7). False alarm rates did not differ significantly between groups (t[13] = 0.26, BF = 0.28), nor did the proportion of trials in which more than one face was reported (t[13] = 0.77, BF = 0.38). At the neural level, we found that EEG amplitude co-varied with stimulus intensity and detection irrespective of STN stimulation on a cluster of electrodes around CP1, from 0.38 to 0.56 ms after stimulus presentation. Follow-up Bayesian analyses at the cluster-lever (model 5) confirmed the interaction between intensity and detection (estimate 2.65 [1.66 3.61], BF > 6000). However, they revealed no reliable evidence for a main or interaction effect of stimulation (Fig. 4C-D, SI Table 8-9).

**Figure 4.**
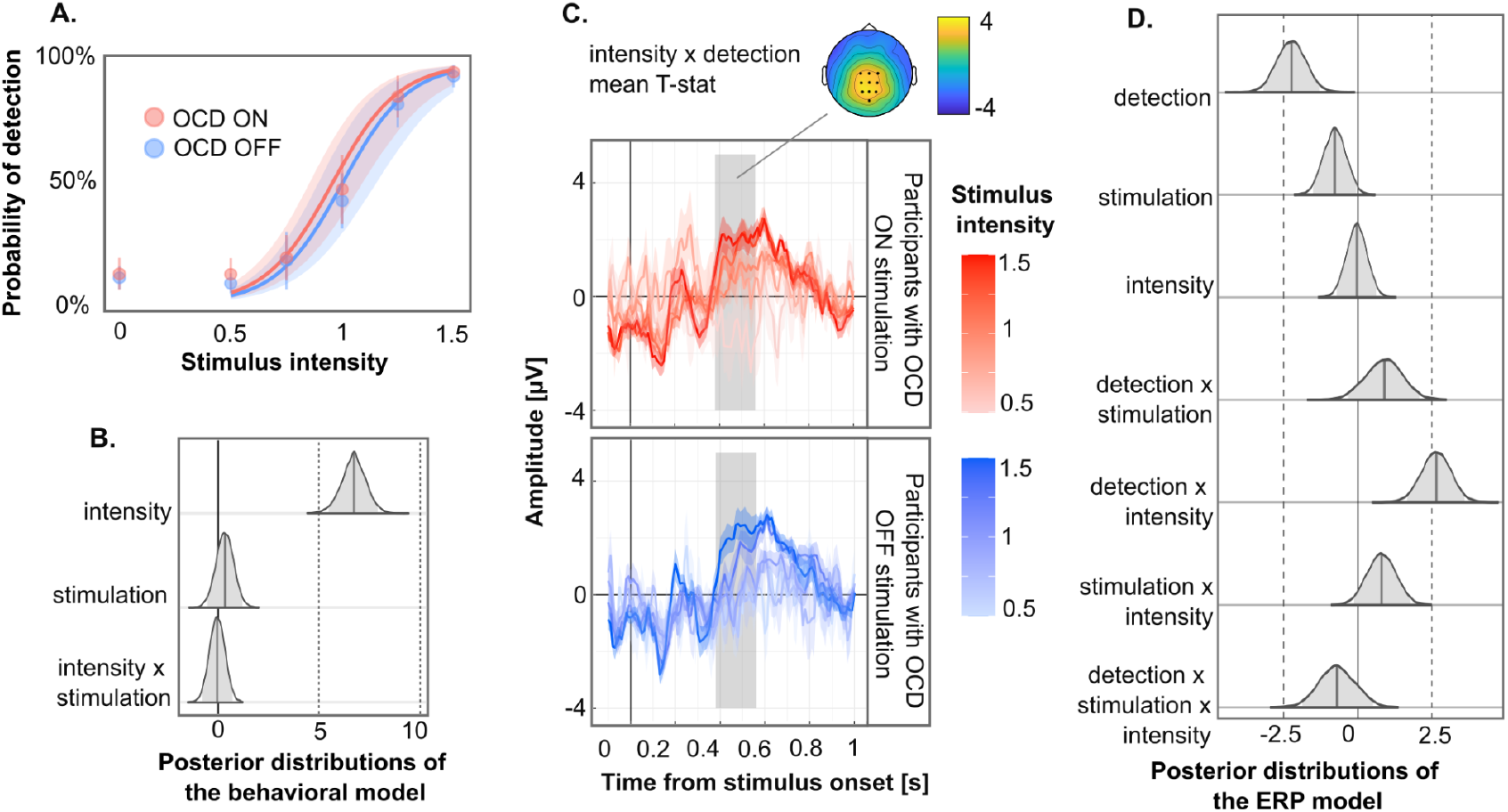
Participants with OCD off-stimulation (in blue) and on-stimulation (in red). **A. Detection reports.** The dots show the mean probability of detection for each stimulus intensity level. The line and ribbon show the Bayesian model prediction and 95% confidence interval. **B. Posterior distributions from the Bayesian model predicting detection reports. C. Detection-related evoked potential**. The topography represents the mean t-statistic of the most significant effect during the time window, i.e., the interaction between intensity and detection on CP1 (190ms) from 0.38 to 0.56 ms after stimulus presentation. The dots represent the electrodes of interest, i.e., electrodes responding to intensity for more than 50 ms in a row, within the time window of interest, and surrounding the most responsive electrode, P1. Below is the amplitude of the evoked ERP on the cluster of interest. The time window of interest is shown in grey. **D. Posterior distributions from the Bayesian model predicting EEG amplitudes depending on detection reports**.

In sum, DBS did not alter stimulus detectability. Next, we checked if DBS modulated confidence and possibly alleviated the metacognitive deficits we observed among patients. To do so, we examined confidence ratings depending on the intensity of detected and undetected stimuli in patients on- and off-stimulation. Contrary to the control-patients analysis, and against what we had pre-registered, we found evidence in favour of an absence of an interaction between STN stimulation and intensity (estimate =0.08 [−0.38 0.54], BF = 0.12), detection (estimate = 0.26 [−0.49 1.02], BF = 0.25) or intensity and detection (estimate = 0.06 [−0.68 0.80], BF = 0.18; Fig. 5A-B, SI table 13). At the neural level, we found significant EEG amplitude modulations by stimulus intensity and confidence over only one occipital scalp location (PO8). Follow-up Bayesian analyses at this location revealed evidence in favour of an interaction between stimulation and confidence (estimate= −2.36 [−4.85 0.1], BF = 3.57), irrespective of stimulus intensity (Figure 5C-D and SI tables 14-15). Together, these results indicate that subthalamic stimulation affected one cortical evoked potential related to confidence but failed to alleviate the metacognitive deficit identified among patients compared to control participants.

**Figure 5.**
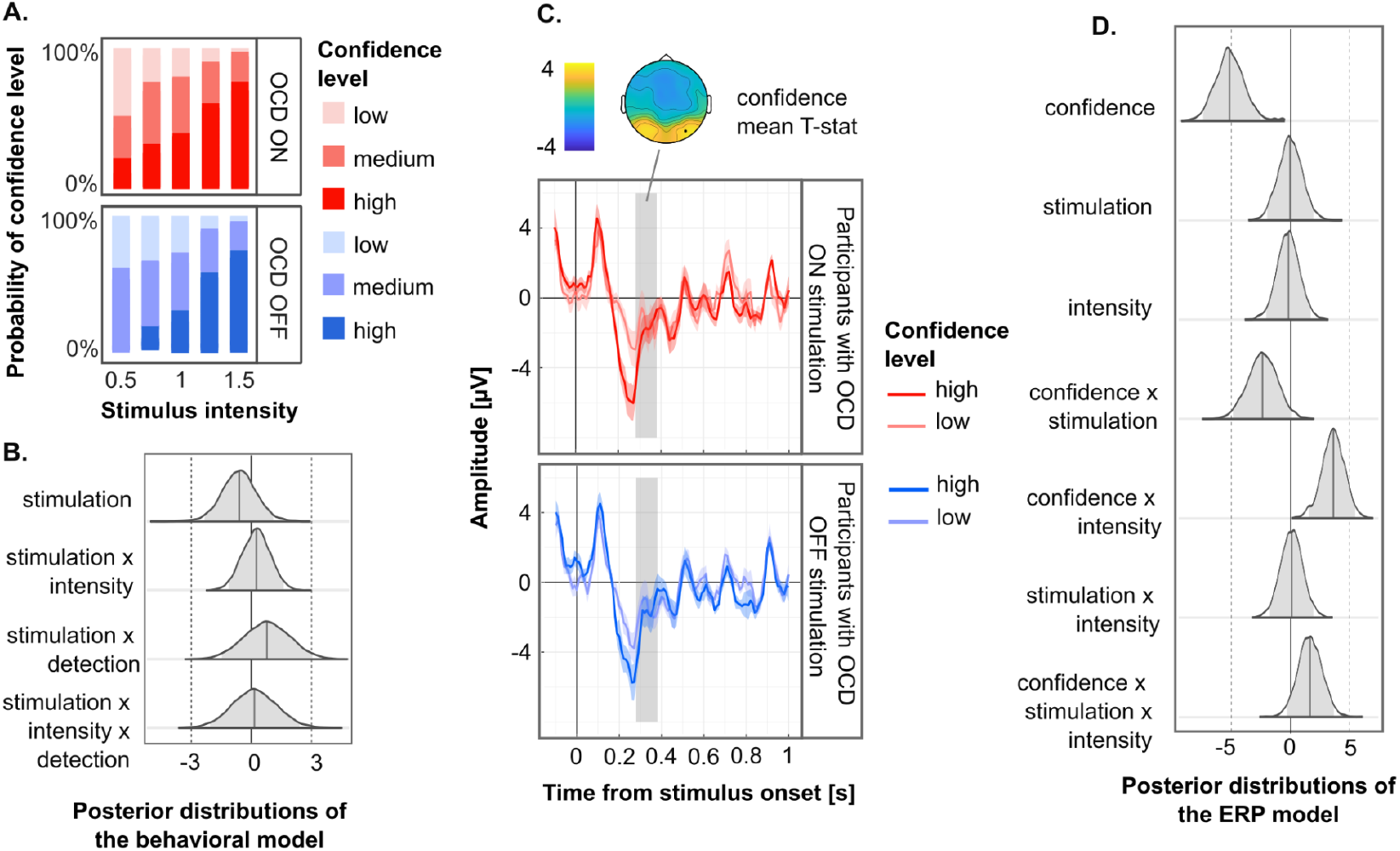
Participants with OCD off-stimulation (in blue) and on-stimulation (in red). **A. Confidence reports**. Probability of confidence level for each stimulus intensity level. **B. Posterior distributions from the Bayesian model predicting confidence reports**. Only effects related to the subthalamic stimulation are shown here. **C. Confidence-related evoked potential**. The topography represents the mean t-statistic of the most significant effect during the time window, i.e., the main effect of confidence, between 0.28 and 0.38 after stimulus presentation. The dots represent the electrode of interest, i.e., PO8 electrode responding to intensity for more than 50 ms in a row, within the time window of interest. Below is the amplitude of the evoked ERP on the cluster of interest. The time window of interest is shown in grey. Low and medium-confidence reports have been pooled into a single Low level to increase plot readability, and all stimulus intensity levels are pooled together. **D. Posterior distributions from the Bayesian model predicting EEG amplitudes depending on confidence reports**.

## Discussion

We used a visual detection and confidence judgement task to assess perceptual and metacognitive performances in OCD. We found that individuals with OCD have preserved visual detection abilities but impaired metacognitive performance (when off stimulation) compared to control participants. This result suggests a specific impairment at the metacognitive level, which cannot be explained by a defect in first-order perceptual processing as is commonly the case in psychiatric disorders (Hoven et al., 2019; Rouy et al., 2021). Contrary to our hypothesis, we found no lowering of the detection threshold nor compensation for the metacognitive deficit under STN stimulation.

We hypothesised two behavioural differences between the OCD and control groups. First, contrary to our hypothesis, we found no evidence for better stimulus detectability among control participants. Second, we confirmed the existence of a metacognitive deficit that was previously found in individuals with sub-clinical OCD traits such as high compulsivity (Hauser et al., 2017), compulsive behaviour, and intrusive thought (Rouault et al., 2018). This result contrasts with a recent study which showed — with a different methodology — that the metacognitive deficit found in highly compulsive controls was not present in individuals with clinically relevant OCD (Hoven et al., 2023). Exploratory analyses in our study revealed no over-or under-confidence in the OCD group, which is found in some but not all memory studies (Hoven et al., 2019). Interestingly, we found evidence against a corresponding scaling of the ERP correlate of detection over the centroparietal cortex, previously linked to evidence accumulation and confidence (O’Connell et al., 2012; Pereira et al., 2021). This absence of effect suggests that the neural locus of this putative metacognitive deficit could be downstream, in frontal regions that are commonly associated with OCD. Although this result is clinically relevant and can contribute to better management of OCD, the comparison of a population with psychiatric disorder with a so-called control population is undermined by numerous pitfalls (Schechter et al., 1998). Among others, the comparison with a healthy population is too unspecific to study precise, circumscribed clinical dimensions. Here, we took advantage of the fact that patients were treated with subthalamic DBS to evaluate - in a within-subject design - the role of the STN in perception and metacognition. Contrary to what we had preregistered, we found evidence in favour of the absence of an effect of subthalamic DBS on both visual detection and confidence.

The causal effect of the STN on decision threshold was observed previously during immediate response-tasks with dorsal stimulation in Parkinson’s disease (Cavanagh et al., 2011; Frank et al., 2007), but also ventromedial stimulation in OCD (Kibleur et al., 2016; Voon et al., 2017). The reason why we found no effect may be that the STN modulates decision thresholds for immediate, but not delayed responses such as in our paradigm. This possibility, however, seems at odds with the fast response of STN neurons to sensory and perceptual content even in the absence of an immediate response (Pereira et al., 2024) or the fact that the effect STN-DBS on response times is restrained to a short period post-stimulus (Herz et al., 2018). Another possibility is that STN-DBS mediates the inhibition between two competing response accumulators (Green et al., 2013). In the case of a detection task with only one accumulator, the STN would have no causal effect on the decision. There may also be several other reasons why subthalamic DBS was ineffective in modulating decision thresholds or restoring metacognitive performance in our study. We note that the effectiveness of STN DBS on OCD is not in doubt (Chabardes et al., 2020) since the symptoms were drastically lower when participants were enrolled in the research protocol (mean YBOCS: 13.4) compared to before DBS surgery (mean YBOCS: 33.4). However, symptoms before and after the 16h wash-out period within our experimental protocol were similar, suggesting that not all cognitive processes relevant to OCD symptomatology are affected by acute changes in subthalamic stimulation. Indeed, while anxiety is rapidly influenced by stimulation, mood changes, and OCD symptoms can take much longer to be affected by stimulation (Ashkan et al., 2017). Therefore, this wash-out period may be insufficient for observing behavioural changes in our task. Extending the washout period is disputable, given the discomfort it imposes on patients. Notably, this wash-out period was long enough to observe non-specific side effects that may have precluded the expected effects, including fatigue and headaches, consistent with the literature about STN-DBS for Parkinson’s Disease (Bjerknes et al., 2020).

Finally, the variability in stimulation parameters might also have contributed to the inconsistent effects observed across individuals (see SI table 3). Considering this interindividual variability and linking metacognitive deficits with symptomatology requires working with larger cohorts of patients. This remains a significant challenge today, given that subthalamic DBS remains an experimental treatment with few beneficiaries for OCD (Visser-Vandewalle et al., 2022). Nevertheless, the rise of DBS in neurology and psychiatry portends better clinical management of metacognitive deficits and a window to causally explore the thalamocortical circuits involved in decision-making and metacognition.

## Contributions

Michael Pereira and Nathan Faivre designed the study. Audrey Kist, Michael Pereira, Ramla Msheik, Dorian Goueytes, Arnaud Pouchon collected the data. Michael Pereira, Audrey Kist, and Nathan Faivre analysed the data. Stephan Chabardès performed DBS surgeries. Mircea Polosan supervised all clinical aspects of the study. Audrey Kist, Michael Pereira, and Nathan Faivre wrote the article, which all authors reviewed. All authors approved the final version of the manuscript. This work was presented at the Association for the Scientific Study of Consciousness in Amsterdam in July 2022.

## Data availability

The experimental procedure and analysis plan have been preregistered (https://osf.io/qcvsd). All analysis scripts are publicly available (https://gitlab.com/nfaivre/ocd_dbs_public). Data will be made publicly available upon publication.

## Acknowledgements

We thank Margot Pourady for her help with Figure 1.

