## Supplementary information for "Obsessive-compulsive disorder is characterised by metacognitive deficits resistant to subthalamic stimulation"

**Affiliations**

#### Participants

We recruited participants who met DSM-IV criteria for severe, resistant OCD and agreed to participate in a clinical trial on DBS at the Grenoble Alps University Hospital (NCT 02844049). The protocol was approved by the Ethics Committee CPP Sud-Est V. The sample size was based on the available cohort of patients with OCD in the STN DBS clinical trial at Grenoble University Hospital, which was consistent with previous studies.

The present work is independent of the clinical trial, and none of its pre-specified primary outcomes is mentioned in the present manuscript. All participants reported being right-handed except for one left-handed participant. All participants had normal or corrected-to-normal vision except for one with congenital one-eyed blindness, showing detection performances similar to other participants. Three patients were excluded from behavioural analyses: two who interrupted the experiment before completion and one with a false-alarm rate of 49.7%, suggesting that instructions were not followed accurately. One control participant was excluded because of a technical problem. One additional patient was excluded from EEG analyses due to a technical problem.

| | OCD (baseline, n=14) | Control (n=18) | t-statistic | BF |
|---|---|---|---|---|
| Age, yr | 44.07 ± 9.99 | 45.5 ± 11.71 | -0.38 | 0.36 |
| Education, yr | 14.79 ± 2.66 | 16 ± 2.53 | -1.35 | 0.68 |
| ACE-r | 92.9 ± 6.07 | 95.83 ± 3.17 | -1.47 | 1.09 |
| BAI | 6.57 ± 4.24 | 3.65 ± 4.42 | 1.93 | 1.35 |
| BDI-FS | 2.86 ± 3.13 | 1.71 ± 1.7 | 1.20 | 0.61 |

**SI Table 1 - Clinical and neuropsychological characteristics of control participants and patients with OCD**. ACE-r = Adenbrook Cognitive Examination revised, BAI = Beck Anxiety Inventory, BDI = Beck Depression Inventory Fast Screen, yr = year, M = mean.

| | Off DBS | On DBS | t-stat | BF |
|---|---|---|---|---|
| Y-BOCS-R | 3.38 | 2.46 | 1.8 | 0.99 |
| Y-BOCS C | 3.92 | 3.23 | 1.6 | 0.74 |
| BAI | 5.77 | 4.23 | 1.1 | 0.46 |
| BDI-FS | 1.92 | 2.15 | -0.8 | 0.36 |

**SI Table 2. Clinical characteristics of participants with OCD**. Y-BOCS = Yale-Brown Obsessive-Compulsive Scale; Y-BOCS R = Y-BOCS Resistance Item; Y-BOCS C = Y-BOCS Control Item; BAI = Beck Anxiety Inventory, BDI-FS = Beck Depression Inventory Fast Screen.

| Participant | Sex | Age range | Education (years) | Disease duration before surgery (years) | Stimulation characteristics | Stimulation duration (years) | Y-BOCS baseline | Y-BOCS on recording | Treatment on recording |
|---|---|---|---|---|---|---|---|---|---|
| 1 | M | 56-60 | 12 | 18 | 2.5 mA - 80 Hz | 17 | 37 | 17 | Lorazepam, Floxyfral, Formoterol |
| 2 | F | 51-55 | 15 | 15 | 4.8 mA - 70Hz - 60 µs / 5.5 mA - 70Hz - 60 µs | 11 | 36 | 18 | Fluoxetine, Hydroxyzine, Pentaprazole |
| 3 | F | 56-60 | 12 | 20 | 3.3 mA - 130 Hz - 30 µs | 2 | 32 | 11 | Fluoxetine, Quetiapine, Prazepam |
| 4 | F | 36-40 | 17 | 21 | 2.2 mA - 70 Hz - 60 µs | 6 | 30 | 21 | Venlafaxine |
| 5 | F | 41-45 | 12 | 18 | 1.7 mA -130 Hz - 60 µs/ 3.0 mA - 130 Hz - 60 µs | 7 | 30 | 1 | None |
| 6 | F | 26-30 | 12 | 17 | 1.8mA - 130Hz - 60 µs / 1.6mA - 130 Hz - 60 µs | 2 | 31 | 20 | Clomipramine, lamotrigine, Isotretinoin |
| 7 | M | 46-50 | 17 | 29 | 1.7mA - 130 Hz - 30 µs | 1 | 32 | 10 | Clomipramine, Fluoxetine, Omeprazole |

| Participant | Sex | Age range | Education (years) | Disease duration before surgery (years) | Stimulation characteristics | Stimulation duration (years) | Y-BOCS baseline | Y-BOCS on recording | Treatment on recording |
|---|---|---|---|---|---|---|---|---|---|
| 8 | M | 31-35 | 15 | 20 | 2.8mA - 130 Hz - 30 µs | 2 | 31 | 13 | Clomipramine, Fluoxetine, Aripiprazole, Oxazepam, Tadalafil |
| 9 | F | 46-50 | 20 | 11 | 1.8 mA - 130 Hz - 60 µs | 12 | 36 | 8 | None |
| 10 | F | 36-40 | 15 | 24 | 2.6 mA - 130 Hz - 60µs | 9 | 34 | 20 | Levothyroxine |
| 11 | F | 36-40 | 12 | 25 | 1.3 mA - 130 Hz - 60µs | 4 | 32 | 14 | Zoloft, Sulfarlem, Esomeprazole, Clomipramine |
| 12 | F | 46-50 | 15 | 30 | 4.1 mA - 130 Hz - 30 µs / 4.0 mA - 130 Hz - 30 µs | 5 | 34 | 6 | Fluoxetine, Levothyroxine |
| 13 | F | 56-60 | 18 | 20 | 1. 7 mA - 130 Hz - 30 µs | 1 | 35 | 12 | Fluoxetine, Clomipramine, Haloperidol, Levothyroxine, Bromazepam, Melatonin |
| 14 | F | 30-35 | 15 | 15 | 1.7mA - 130 Hz - 30 µs | 1 | 37 | 11 | None |

**SI Table 3 - Clinical characteristics of participants with OCD.** Stimulation characteristics are reported for the left and right STN unless stimulation parameters are the same on both sides.

#### Neuropsychological and clinical evaluation

We assessed cognitive ability with the Addenbrooke Cognitive Examination Revised – ACE-r[1] among control participants at the time of their inclusion in this experiment and among individuals with OCD at the time of their inclusion in the DBS therapy research protocol. We assessed anxiety and depression symptoms using the Beck Anxiety Inventory - BAI[2] and the Beck Depression Inventory Fast Screen - BDI-FS[3] in control participants, participants with OCD on-stimulation, and participants with OCD off-stimulation. Moreover, we assessed the long-term effects of STN stimulation on OCD symptoms with the Yale-Brown Obsessive Compulsive Scale - Y-BOCS[4,5] before patients received the stimulation device and on stimulation at the time of the experiment. We also assessed rapid changes in symptomatology by comparing the Y-BOCS items Resistance (R) and Control (C) on and off stimulation.

#### Procedure

Face stimuli embedded in a sequence of noise frames were presented using the Psychophysics toolbox version 3[6–8] in Matlab (Mathworks) for Ubuntu 18.04.4 LTS. Participants were installed at approximately 60 cm from a laptop screen (Dell Precision 7530, 17 inches, 1920*1080 px, 60 Hz, maximum luminosity). After a fixation cross (random duration: 0.25 - 0.75 s), participants were presented with a 4 s sequence of 20 grayscale images of visual noise (i.e. one every 200 ms) at a visual angle of 4.8° (Figure 1). Each noise image was obtained by randomly shuffling the phase of the face stimulus. A face stimulus was superimposed on one of the 20 visual noise frames at a random onset ranging between 0.5 and 3.5 s after the beginning of the sequence. Stimulus intensity was manipulated by changing the contrast between the face and the noise frame (0, 0.5, 0.75, 1, 1.25, or 1.5 times each participant's detection threshold, measured before the experiment; see below). At the end of the sequence, following a delay ranging from 0.25 to 0.75 s, participants were asked to report the number of faces they had detected by pressing a key with their right hand corresponding to "zero", "one", or "more than one". Of note, no more than one face was ever presented per trial. This response option was added based on pilot participants reporting seeing two faces in a small fraction of trials. Participants were then prompted to report their confidence in their answer on a 3-level scale using another set of buttons on the keypad, corresponding to: "not sure", "moderately sure", and "very sure". Individual detection thresholds were estimated before the experiments using a 30-trial one-up/one-down staircase procedure[9], starting with a contrast of 0.15 and with a step size of 5 %. The resulting perceptual threshold was then held constant during all experimental blocks. In the group of participants with OCD, threshold was estimated on-stimulation.

#### Behavioural analysis

All analyses were performed using R version 4.0.3[10] and RStudio 2022.02.3+492[11] , including the tidyverse[12], brms[13] and sJplot packages[14]. Trials in which participants reported seeing more than 1 face (mean 8.2%± 15.4% of trials per participant) and trials with extreme reaction times on the detection report (i.e., 2.5% slowest and 2.5% fastest responses) were excluded. Evidence in favour of the alternative hypothesis is considered moderate if BF > 3. It is considered weak if BF < 3 but with a 95% credible interval not overlapping with zero. Evidence in favour of the null hypothesis is considered moderate if BF < 1/3.

Model 1 contained a random intercept for participants and experimental blocks nested within each participant. We specified priors for fixed effects as follows:

- As we expected a positive effect of the stimulus intensity, we set a mildly informative prior with a normal distribution of mean = 3 and standard deviation = 2: N(3, 2).
- As we expected control participants to have a lower detection threshold than participants with OCD off-stimulation, and patients off-stimulation to have a lower detection threshold than patients on-stimulation, we set a mildly informative prior N(1,2) for the main effect of the group. We also expected a possible interaction between intensity and group, and accordingly set a prior N(1, 2).
- As we expected no other main or interaction effect, we set zero-centered priors N(0, 2) for other fixed effects. All other priors followed the default option in the brms package.

Model 2 contained a random intercept for participants and blocks nested within participants. We used a N(3, 2) prior for the effect of detection, and a mildly informative prior N(1, 2) for the main and interaction effects of the group. All other priors followed the default option in the brms package.

#### EEG preprocessing

EEG was recorded with an actiCHamp amplifier (Acticap 10-10 layout; Brain Products, Inc., Gilching, DE). Horizontal electrooculography (hEOG) was recorded using a bipolar montage consisting of two electrodes placed on the outer canthi of each eye. Vertical electrooculography (vEOG) was recorded using a bipolar montage with left supra- and infra-orbital electrodes. For participants in the OCD group, we recorded the DBS signal by placing two additional electrodes at the base of the neck on both sides of the subcutaneous wire connecting the stimulator in the chest to the DBS electrodes. For all auxiliary bipolar pairs, ground electrodes were installed on the left shoulder blade. The EEG signal was first lowpass filtered with a 45 Hz cutoff two-pass Butterworth filter to remove both the line noise at 50 Hz and the DBS artifact (130 Hz for all patients except one at 70 Hz and one at 80 Hz, see Supplementary Table 3 for details). The signal was then downsampled to 500 Hz and highpass filtered with a 0.5 Hz cutoff two-pass Butterworth filter. Finally, a 50 Hz notch filter was applied to remove residual line noise artefacts, and a stimulation subharmonics frequency filter was also applied at half of the stimulation frequency. EEG data were epoched around stimulus onset using a photodiode (except for 6 sessions (21.4 %) in which the photodiode was faulty and replaced by serial port triggers). Defective channels or trials with strong artifacts were rejected after visual inspection. These rejected channels were replaced with the average of all neighbouring channels as obtained by triangulation from a 2-dimensional projection of the sensor positions with the Delaunay method. Finally, data were re-referenced to a common average. To remove ocular, myographic, and any remaining DBS artefacts, we performed an Independent Components (IC) Analysis[15] using FastICA[16] while retaining 99% of the variance from the electrode space[17]. IC activations were correlated to horizontal and vertical EOG and DBS artefacts and removed if the Pearson correlation coefficient was higher than 0.2. Additional ICs were manually removed if deemed artifactual after visually examining their time, frequency and topographic representations. These procedures led to an average of 29.6 ±2.2 independent components (ICs) for participants with OCD off-stimulation, 29.64 ±1.8 ICs for participants with OCD on-stimulation, and 33.2 ±2.0 ICs for control participants.

#### EEG analysis

As trials with confidence rated as “not sure” were rare (10,9%), we pooled them with “moderately sure” reports (28.0%) and compared them to “very sure” reports (61.1%). Electrodes were considered responsive when one of the parameters had a significant effect for at least 50 ms after adjusting for false discovery rate.

We specified priors as follows:

- We expected higher voltage amplitudes following detected versus undetected faces and accordingly set a mildly informative Gaussian prior N(1,2) for the main effect of detection. We also expected higher amplitudes following high-intensity stimuli and set a Gaussian prior N(1,2) for the main effect of intensity. Finally, we expected ERP amplitudes to be higher for perceived than unperceived stimuli and accordingly set a Gaussian prior N(1;2) for the interaction between intensity and response
- We expected higher voltage amplitude in control participants compared to patients, and in patients on-stimulation compared to off-stimulation. Accordingly, we set a default normal distribution centred on (1, 2) for the group parameter.
- We expected no other main or interaction effect, so we set a default normal distribution centred on zero (0, 2) for all other fixed effects. All other priors followed the default option in the brms package.

All Bayesian models were created in Stan computational framework (http://mc-stan.org/) accessed with the brms package, based on four chains of 4000 iterations, including 1000 warmup samples for each chain. All R-hat values were checked to be close to 1, indicating convergence of the models. Bayes factors were computed using the Savage-Dickey method. We interpreted a Bayes factor (BF) > 3 as providing moderate evidence in favour of the tested hypothesis and a Bayes Factor < 0.3 in favour of the null hypothesis. We interpreted a Bayes factor (BF) > 5 as providing strong evidence in favour of the tested hypothesis and a Bayes Factor < 0.2 in favour of the null hypothesis. A Bayes Factor between 0.3 and 3 is considered inconclusive, unless the 95% of the corresponding posterior distribution does not overlap with zero (i.e., in that case it is considered weak evidence).

#### Electrodes reconstruction

All patients had previously undergone preoperative T1 MRI and postoperative CT scans, which were used to reconstruct electrode positions in the STN.

For participants with OCD undergoing STN DBS, we used the Lead-DBS image reconstruction Matlab toolkit[18,19] to coregister preoperative MRI with post-operative CT scans. We used advanced normalisation tools ANTs[20,21] to co-register preoperative T1-weighted 3D images with postoperative CT, including a brainshift correction using the default option 'coarse mask'[22]. Then, based on the preoperative volumes, we used the non-linear segmentation method[23] of the Statistical Parametric Mapping SPM12[24] to compute a multispectral normalisation in the asymmetric nonlinear space ICBM 2009b[25]. Then, the PaCER method was applied to automatically reconstruct the DBS electrodes and their directional contacts[26]. To determine the location of each electrode contact relative to STN subdivisions, we segmented the reconstructed images using the DISTAL atlas[27].

### Detection analysis - detailed results

#### Participants with OCD on-subthalamic stimulation compared to control participants

| **Effect** | **estimate** | **CI** | **R(hat)** | **BF** | **evidence** |
|---|---|---|---|---|---|
| Group | -0.37 | [-1.89 1.14] | 1.00 | 0.38 | Inconclusive |
| Intensity | 6.63 | [5.46 7.79] | 1.00 | <8000 | Strong, in favour of H1 |
| Group * Intensity | 0.56 | [-0.94 2.02] | 1.00 | 0.41 | Inconclusive |

**SI Table 4 - Detailed behavioural results of stimulus detectability (patients off DBS vs controls).** Model 1: detection ~ group * intensity + onset + (group + intensity | participant/block).

False alarm rates did not significantly differ between groups (t[13] = 0.87, BF = 0.50), nor did the proportion of trials in which more than one face was reported (t[13] = 1.43, BF = 0.85).

| Model parameter (Off-ctr) | Responsive electrodes |
|---|---|
| Intensity | O1, O2, Oz, P5, P7, PO3, PO4, PO7 |
| Response | C1, C2, CP1, CP2, CPz, Cz, FCz, FT10, FT9, O1, O2, Oz, P1, P2, P8, PO4, PO7, PO8, Pz, TP10 |
| Group | None |
| Intensity * detection | C1, C2, CP1,CP2, CP3, CPz, Cz, FC1, FCz, FT10, FT9, Fz, O1, O2, Oz, P1, P2, P6, P8, PO3, PO4, PO7, PO8, POz, Pz, TP10 |
| Intensity * group | FCz |
| Group * detection | None |
| Intensity * group * detection | Oz |
| **Longest effect** | Intensity * detection on CPz (290 ms) from 0.38 to 0.66 s |
| **Cluster of interest during longest effect** | C1, C2, CP1, CP2, CP3, CPz, Cz, P1, P2, POz, Pz. |

**SI Table 5 - Selection of electrodes responsive to each parameter of the detection model when comparing participants with OCD off-stimulation to control participants.** Model 3: amplitude ~ intensity * detection * group + (1|participant). The model was fitted separately at each time point, and FDR correction was applied. Electrodes are considered responsive when a parameter has a significant effect during at least 50 ms. The cluster of interest consists of electrodes responding to intensity * detection for more than 50 ms in a row, between 0.38 and 0.66 after stimulus presentation and surrounding most responsive electrode CPz. Smaller effects were found, including an interaction between stimulus intensity and group between 0.42 and 0.46 on FCz, and an interaction between stimulus intensity, group, and detection on Oz between 0.91 and 0.95 ms. These effects were not analysed further, given their brief and focal nature.

| Effect | estimate | CI | Rhat | BF | evidence |
|---|---|---|---|---|---|
| Detection | -2.01 | [-2.67 -1.34] | 1.00 | > 16000 | Strong in favour of H1 |
| Group | -0.46 | [-0.93 0.01] | 1.00 | 1.59 | Inconclusive |
| Intensity | 0.09 | [-0.37 0.56] | 1.00 | 0.12 | Strong in favour of H0 |
| Detection * Group | 0.59 | [-0.33 1.48] | 1.00 | 0.48 | Inconclusive |
| Detection * Intensity | 2.57 | [1.91 3.22] | 1.00 | > 16000 | Strong in favour of H1 |
| Group * Intensity | 0.59 | [-0.03 1.22] | 1.00 | 0.85 | Inconclusive |
| Detection * Group * Intensity | -0.20 | [-1.13 0.75] | 1.00 | 0.23 | Moderate in favour of H0 |

**SI Table 6 - Detailed results of the Bayesian model of evoked potential related to detection on the selected electrodes cluster and time window of interest when comparing participants with OCD off-stimulation to control participants**
Model 5: amplitude ~ detection * group * intensity + (detection * group * intensity | participant)

#### Participants with OCD on-subthalamic stimulation compared to off-stimulation

| Effect | estimate | CI | Rhat | BF | evidence |
|---|---|---|---|---|---|
| Stimulation | 0.34 | [-0.54 1.21] | 1.00 | 0.25 | Moderate in favour of H0 |
| Intensity | 6.72 | [5.48 7.91] | 1.00 | > 16000 | Strong evidence in favour of H1 |
| Stimulation * Intensity | -0.04 | [-0.80 0.72] | 1.00 | 0.18 | Strong in favour of H0 |

**SI Table 7 - Detailed results of OFF-ON Bayesian model of detection behaviour.** Model 1: detection ~ stimulation * intensity + onset + (stimulation + intensity | participant/block)

| Model parameter | Responsive electrodes in Off-on comparison |
|---|---|
| Intensity | O1, O2 (70ms), Oz (70 ms), P7, PO3, PO4, PO7 |
| Response | CP1, CP2, Cz, O1, O2, Oz, P1, P8, PO4 (80ms), PO7, PO8, Pz, TP10, TP9 |
| Group | None |
| Intensity * Detection | C1, C2, CP1 (190 ms), CP2, CPz, Cz, FC1, FT9, O1, O2, Oz, P1, P2, P6, P8, PO3, PO4, PO7, PO8, POz, Pz, TP10 |
| Intensity * Stimulation | None |
| Stimulation * Detection | None |
| Intensity * Stimulation * Detection | None |
| Longest effect | Intensity * detection on CP1 (190ms) from 0.38 to 0.56 ms after stimulus presentation |
| Cluster of interest during longest effect | C1, C2, CP1, CP2, CPz, Cz, P1, P2, POz, Pz |

**SI Table 8 - Selection of electrodes responsive to each parameter of the detection model when comparing participants with OCD off-stimulation to on-stimulation.** Model 3: amplitude ~ intensity * detection * stimulation + (1|participant). The model was fitted separately on each timepoint and each electrode, and an FDR correction was applied. Electrodes were considered responsive when a parameter had a significant effect for at least 50 ms in a row. Therefore, the cluster of interest consists of electrodes responding to intensity * detection for more than 50 ms in a row, between 0.38 and 0.56 after stimulus presentation, and surrounding the most responsive electrode CP1.

| Effect | estimate | CI | Rhat | BF | evidence |
|---|---|---|---|---|---|
| Detection | -2.24 | [-3.18 -1.29] | 1.00 | >16000 | Strong in favour of H1 |
| Stimulation | -0.78 | [-1.51 -0.05] | 1.00 | 1.39 | Weak in favour of H1 |
| Intensity | -0.03 | [-0.71 0.63] | 1.00 | 0.15 | Strong in favour of H0 |
| Detection * Stimulation | 0.89 | [-0.33 2.08] | 1.00 | 0.70 | Inconclusive |
| Detection * Intensity | 2.65 | [1.66 3.61] | 1.00 | >16000 | Strong in favour of H1 |
| Stimulation * Intensity | 0.81 | [-0.13 1.77] | 1.00 | 0.81 | Inconclusive |
| Detection * Stimulation * Intensity | -0.69 | [-1.87 0.51] | 1.00 | 0.49 | Inconclusive |

**SI Table 9 - Detailed results of the Bayesian model of evoked potential related to detection on the selected electrode cluster and time window of interest when comparing participants with OCD off-stimulation to on-stimulation.**
Model 5: signal amplitude ~ detection * group * intensity + (detection * group * intensity | participant).

### Confidence analysis - detailed results

#### Participants with OCD compared to control participants

| Effect | estimate | CI | Rhat | BF | evidence |
|---|---|---|---|---|---|
| Group | 0.14 | [-0.82 1.10] | 1.00 | 0.24 | Moderate in favour of H0 |
| Intensity | -0.14 | [-0.49 0.21] | 1.00 | 0.11 | Strong in favour of H0 |
| Detection | -4.05 | [-4.97 -3.11] | 1.00 | > 16000 | Strong in favour of H1 |
| Group * Intensity | -0.04 | [-0.51 0.43] | 1.00 | 0.12 | Strong in favour of H0 |
| Group * Detection | -0.87 | [-2.09 0.35] | 1.00 | 0.65 | Inconclusive |
| Intensity * Detection | 3.73 | [3.18 4.28] | 1.00 | > 16 000 | Strong in favour of H1 |
| Group * Intensity * detection | 0.90 | [0.14 1.65] | 1.00 | 2.56 | Weak, in favour of H1 |

**SI Table 10 - Detailed results of OFF-CTR Bayesian model of confidence behaviour.** Model 2: confidence ~ group * intensity * detection + onset + (group + intensity + detection | participant/block).

| Model parameter | Responsive electrodes in Off-ctr comparison |
|---|---|
| Intensity | C1, C2, CP1, CP2, CPz, Cz, FCz, FT10, O1, O2, Oz, P1, P2, P5, P6, P7, P8, PO3, PO4, PO7, PO8, POz, Pz, TP10 |
| Confidence | P5, PO3, PO7, PO8 |
| Group | None |
| Intensity * Confidence | P5, PO3, PO8 |
| Intensity * group | None |
| Group * Confidence | None |
| Intensity * group * confidence | None |
| Longest effect | Intensity on Pz (140ms) from 0.38 to 0.51 |
| Cluster of interest | C1, CP1, CP2, CPz, Cz, P1, P2, PO3, Pz |

**SI table 11 - Selection of electrodes responsive to each parameter of the confidence model when comparing participants with OCD off-stimulation to control participants**. Model 4: amplitude(HIT) ~ intensity * group * confidence + (1|participant). The model was fitted separately at each timepoint. An FDR correction was applied. Electrodes were considered responsive when a parameter had a significant effect during at least 50 ms in a row. Therefore, the cluster of interest consists of electrodes responding to intensity for more than 50 ms in a row, between 0.38 and 0.51 s after stimulus presentation and surrounding the most responsive electrode Pz.

| Effect | estimate | CI | Rhat | BF | evidence |
|---|---|---|---|---|---|
| Group | 0.1 | [-1.38 1.15] | 1.00 | 0.30 | In favour of H0 |
| Intensity | 1.14 | [0.16 2.09] | 1.00 | 2.94 | Weak in favour of H1 |
| Confidence | -2.4 | [-3.85 -0.91] | 1.00 | 100 | Strong in favour of H1 |
| Group * Intensity | 0.63 | [-0.7 1.97] | 1.00 | 0.46 | Inconclusive |
| Group * Confidence | 0.28 | [-1.54 2.03] | 1.00 | 0.41 | Inconclusive |
| Intensity * Confidence | 2.52 | [1.32 3.68] | 1.00 | >8000 | Strong in favour of H1 |
| Group * Intensity * * Confidence | -0.08 | [-1.52 1.38] | 1.00 | 0.33 | Moderate in favour of H0 |

**SI table 12 - Detailed results of the Bayesian model of evoked potential related to confidence on the selected electrodes cluster and time window of interest when comparing participants with OCD off-stimulation to control participants.** Model 6: amplitude(HIT) ~ group * confidence * intensity + (group * confidence * intensity | participant). To increase statistical power, medium-confidence reports have been pooled with low-confidence reports.

**Participants with OCD on-subthalamic stimulation compared to off-stimulation (OFF-ON) Behaviour**

| Effect | estimate | CI | Rhat | BF | evidence |
|---|---|---|---|---|---|
| Stimulation | -0.20 | [-0.77 0.37] | 1.00 | 1.16 | Inconclusive |
| Intensity | -0.02 | [-0.45 0.42] | 1.00 | 0.03 | Strong in favour of H0 |
| Detection | -3.99 | [-5.18 -2.75] | 1.00 | > 16000 | Strong in favour of H1 |
| Intensity * Detection | 3.76 | [3.22 4.30] | 1.00 | > 16000 | Strong in favour of H1 |
| Stimulation * Detection | 0.26 | [-0.49 1.02] | 1.00 | 0.25 | Moderate in favour of H0 |
| Stimulation * Intensity | 0.08 | [-0.38 0.54] | 1.00 | 0.12 | Strong in favour of H0 |
| Stimulation * Intensity * Detection | 0.06 | [-0.68 0.80] | 1.00 | 0.18 | Strong in favour of H0 |

**SI Table 13 - Detailed results of OFF-ON Bayesian model of confidence behaviour.** Model 2: confidence ~ stimulation * intensity * detection + onset + (stimulation + intensity + detection | participant/block).

| Model parameter | Responsive electrodes when comparing Off-on |
|---|---|
| Intensity | CP1, CP2 (90ms), CPz, Cz, O1 (90ms), O2, Oz, P2, P5, P6, P7, P8, PO3 (90ms), PO4, PO7 (90ms), PO8, POz, Pz, TP10, TP9 |
| Confidence | PO3 (90ms), PO8 (110ms) |
| Stimulation | None |
| Intensity * confidence | PO3 (80 ms), PO8 |
| Intensity * stimulation | None |
| Stimulation * confidence | None |
| Intensity * stimulation * confidence | none |
| Longest effect | Confidence effect on PO8, 110 ms from 0.28 to 0.38 |
| Electrode of interest | PO8 |

**SI Table 14 - Selection of electrodes responsive to each parameter of the confidence model when comparing participants with OCD off-stimulation to on-stimulation.** Model 4: amplitude (HIT) ~ intensity * stimulation* confidence + (1|participant). The model was fitted separately at each timepoint, and an FDR correction was applied. Electrodes were considered responsive when a parameter had a significant effect for at least 50 ms in a row. Here, the only electrode of interest, i.e., electrode responding to confidence for more than 50 ms in a row, between 0.28 and 0.38 s after stimulus presentation, is PO8.

| Effect | estimate | CI | Rhat | BF | evidence |
|---|---|---|---|---|---|
| Stimulation | -0.01 | [-1.97 2] | 1.00 | 0.43 | Inconclusive |
| Intensity | 0.17 | [-2.05 1.72] | 1.00 | 0.41 | Inconclusive |
| Confidence | -5.1 | [-7.4 -2.52] | 1.00 | > 8000 | Strong in favour of H1 |
| Stimulation * Intensity | 0.11 | [-1.76 1.99] | 1.00 | 0.42 | Inconclusive |
| Stimulation * Confidence | -2.36 | [-4.85 0.1] | 1.00 | 3.57 | In favour of H1 |
| Intensity * Confidence | 3.62 | [1.55 5.49] | 1.00 | > 8000 | Strong in favour of H1 |
| Stimulation * Intensity * Confidence | 1.67 | [-0.3 3.68] | 1.00 | 1.75 | Inconclusive |

**SI table 15**. **Detailed results of the Bayesian model of evoked potential related to confidence on the selected electrode cluster and time window of interest were obtained when comparing participants with OCD off-stimulation to on-stimulation.** Model 6: amplitude (HIT) ~ stimulation * confidence * intensity + (stimulation * confidence * intensity | participant). To increase statistical power, medium-confidence reports have been pooled with low-confidence reports.
